# Improving walking after lumbar spinal stenosis surgery: co-design and single-arm feasibility trial of the STructured Rehabilitation and InDividualised Exercise and Education (STRIDE) programme

**DOI:** 10.64898/2026.03.28.26349602

**Authors:** Suzanne McIlroy, Lindsay M Bearne, Andrew McCarter, Chloe McPherson, Hema Chaplin, Lisa J Brighton, John Weinman, Sam Norton

## Abstract

**Background:** Lumbar spinal stenosis (LSS) can cause pain and severe walking limitation. Although surgery aims to improve walking, many patients do not achieve clinically meaningful gains. Rehabilitation can improve outcomes, yet existing programmes lack robust evidence and theoretical underpinning. This study aimed to (1) co-design a theory-informed rehabilitation programme to improve walking after LSS surgery, and (2) evaluate feasibility of conducting a future trial and acceptability of the intervention.

**Methods:** A multi-methods study included intervention co-design followed by a single-arm feasibility study. Co-design used an adapted Experience-Based Co-Design approach with patients, carers, and healthcare professionals (n=39), integrating the Behaviour Change Wheel. This resulted in STructured Rehabilitation and InDividualised Exercise and Education (STRIDE), delivered over 12-week pre- and 12-weeks post-surgery, targeting knowledge, expectations, perceived control, physical capability, and fears. Adults aged ≥50 years awaiting LSS surgery were recruited to a before–after feasibility study. Feasibility outcomes included recruitment and retention. Acceptability was assessed using the Theoretical Framework of Acceptability questionnaire (0-5 (high acceptability)) and focus groups. Clinical outcomes measured at baseline, post-prehabilitation, and post-rehabilitation included 6-minute walk distance (6MWD) and mean daily step count over 7 days.

**Results:** Fifteen of 31 eligible participants were recruited (48%; mean age 70 years), with 80% retained to study end (2 decided against surgery, 1 unable to complete final assessment). Acceptability was high (median 5/5, IQR 0). Participants valued the personalised, supportive approach and reported improved motivation and preparation for surgery, though travel was burdensome. Small pre-operative and moderate-to-large post-operative improvements were observed in 6MWD (+49.9 m and +81.6 m) and daily step count (+868 and +1405 steps/day).

**Conclusions:** This co-designed, physiotherapy-led, behaviour-change rehabilitation programme was acceptable to participants, with encouraging recruitment, retention, and signals of improved walking following LSS surgery. The findings support progression to a future trial.

## Introduction

Symptomatic lumbar spinal stenosis (LSS) is a degenerative condition, occurring in 11% of the general population, with incidence increasing with age (1). It significantly restricts walking due to pain, paraesthesia and/or weakness in the legs made worse when in an upright posture (2). Therefore, people with LSS are typically very sedentary (3). Surgery is recommended when symptoms have not substantially improved with conservative management and aims to improve symptoms and walking ability.

Walking is a fundamental component of physical function and healthy aging, linked to diverse physical, psychological, and social benefits (4). Walking is an accessible exercise (5) and is associated with improved function and lower morbidity and mortality in older adults (6). However, 75-90% of people do not increase their walking post-operatively (7, 8), thus remaining at risk of inactivity-related consequences. Understanding why patients do not increase walking after LSS surgery, and developing interventions to support increasing walking and physical activity, is a priority (7).

Walking is a modifiable behaviour (4) and is amenable to behaviour change interventions and rehabilitation (9). Although rehabilitation is recommended for people undergoing surgery for LSS, it is inconsistently provided (10). Post-operative exercise-based or multi-modal rehabilitation reduces pain and disability after lumbar surgery (11–14); however, the evidence is of low to moderate quality, with limited long-term follow-up. Furthermore, research has largely focused on discectomy or fusion, with limited attention to older adults receiving surgery for LSS (12), despite evidence that older adults may respond differently to rehabilitation and behaviour change interventions than younger populations (15). Walking recovery has also been largely neglected as an outcome in existing interventions(16), with only one small-scale feasibility study targeting walking, highlighting the need for further development in this area (16). Consequently, there is a need to prioritise research specifically tailored to the needs and experiences of older people with LSS.

Prehabilitation programmes, aimed at increasing physical and psychological preparation for surgery (17) have demonstrated short term benefits for people undergoing lumbar surgery, such as reduced length of stay (18, 19) and improved pain and physical performance (20, 21). However, these results often attenuate by 6 months post-operatively, suggesting sustained support is needed. Combining prehabilitation with post-operative rehabilitation has the potential to consolidate functional improvements achieved during prehabilitation, reduce long-term post-operative disability, and respond to the complex recovery needs of this population and ultimately increase walking behaviour and physical activity among older adults undergoing surgery for LSS.

To address this challenge, we co-designed a new pre-to post-operative rehabilitation programme aimed at increasing walking behaviour. The Medical Research Council (MRC) recommends using published intervention development approaches when designing complex interventions, to support systematic planning and enhance intervention effectiveness and usability (22). Two complementary approaches are the Behaviour Change Wheel (BCW) (23) and Experience-Based Co-Design (EBCD) (24, 25).

The BCW provides a comprehensive framework for characterising behaviours and designing behaviour change interventions (23, 26). The BCW provides a structured approach to behaviour change, centred on the COM-B model of Capability, Opportunity, and Motivation, and guides intervention design from behavioural diagnosis to targeted strategies (27). The BCW offers limited practical guidance on stakeholder involvement. EBCD addresses this gap by embedding intervention development in the lived experiences of patients, families, and clinicians through structured co-design processes (25), to ensure meaningful, acceptable, and sustainable interventions (27).

Intervention development must assess feasibility and acceptability before moving to effectiveness trials (22). Acceptability reflects how appropriate an intervention is perceived to be by those delivering or receiving it, encompassing emotional, cognitive, and ethical dimensions (28). Assessing acceptability reduces the risk of implementing effective yet impractical or unacceptable interventions (29).

This paper describes the development of the STructured Rehabilitation and InDividualised Exercise and Education (STRIDE) programme, designed to improve walking following surgery for lumbar spinal stenosis, and evaluates the acceptability of the programme in a single-arm feasibility study.

## Materials and methods

This multi-methods study comprised an intervention co-design study using a theoretically informed, user-centred process, followed by a single-arm multi-methods feasibility study. Qualitative focus groups and quantitative measures were used to assess the acceptability of the new rehabilitation programme and to test procedures and estimate parameters for a future trial. The study was conducted at King’s College Hospital, a tertiary centre for spinal surgery in London.

The intervention co-design study is reported using the GUIDance for the rEporting of intervention Development framework (30) and template for intervention description and replication (TIDieR) checklist (31). The feasibility study is reported using CONSORT extension for pilot and feasibility trials (32). To maintain conciseness, a comprehensive description of the co-design study is provided in Supplementary Materials 1, with additional detail provided in the PhD thesis of the first author (33).

Ethical approval was obtained for both studies (East Midlands - Nottingham 1 Research Ethics Committee, 20/EM/0307; London - Bloomsbury Research Ethics Committee, 23/LO/0913), and written informed consent was obtained prior to participation. The feasibility study was pre-registered on ISRCTN: ISRCTN17396524.

### Co-design study

The co-design study was conducted between 29^th^ March and 17^th^ August 2023. It was guided by adapting and integrating principles from EBCD (34, 35) and the BCW (27).

#### Participants

Thirty-nine people participated in the co-design workshops, including 9 patients (who had previously undergone surgery for LSS), 4 family members, and 20 multi-disciplinary healthcare professional (HCP) participants, and 6 facilitators. Patient participants were identified from three NHS hospitals in England after a qualitative interview study exploring experiences of surgery and rehabilitation for LSS (36); eligibility criteria included age ≥50 years, prior surgery for LSS, conversational English language, and no alternative primary cause of pre-operative walking restriction. Clinician participants were purposively sampled to ensure multidisciplinary representation across community and secondary care settings and through professional networks e.g. National Spine Network. The mean age of patient participants was 68 years (range 55-79), 6 (67%) were male, with a range of ethnicities represented (n=13 (46%) White British). Among HCPs, physiotherapists were the largest group (n=12, 60%), followed by spinal surgeons (n=3, 15%); 11 (55%) were female, and 10 (50% were White British).

#### Co-design workshop process

Four, two hour workshops were conducted. The first workshop included only patients and family members, and the second included only HCPs, to facilitate open discussion within each group. The third and fourth workshops involved all participants. Additionally, all participants were invited to a small group meeting (<30 minute duration) in July 2023. Figure 1 illustrates the outputs from the workshops and meetings.

**Figure 1.**
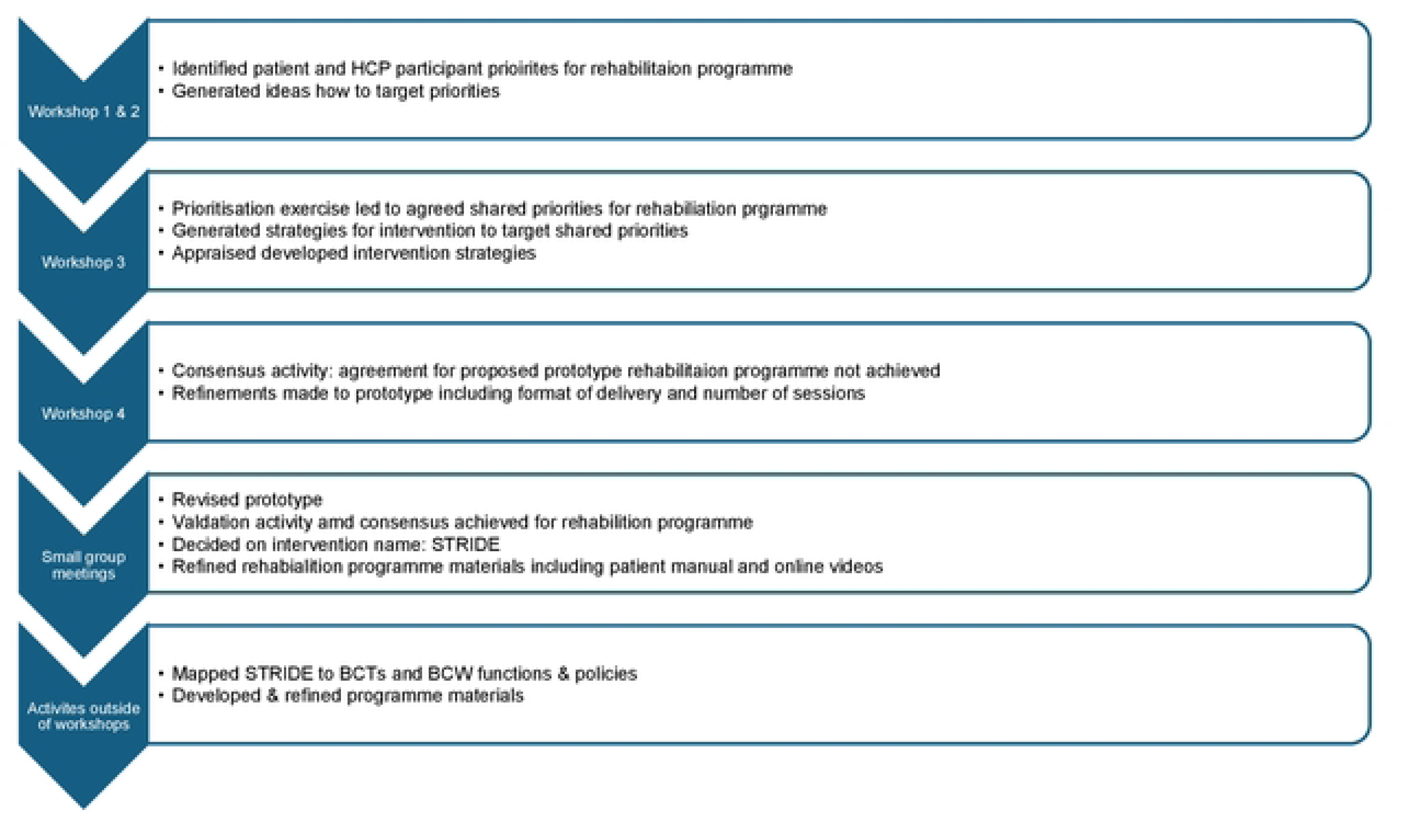
Outputs from co-design workshops and meetings.

#### Workshop one and two

Participants were introduced to the project aims and the problem identified through earlier patient and public involvement, defined behaviourally as: *“People do not increase their walking performance after surgery for LSS.”*

Both workshops aimed to identify preliminary rehabilitation priorities to inform the joint co-design workshop and to generate initial intervention ideas. Key activities included emotional mapping, small-group discussion, and dot voting to explore shared experiences of surgery and recovery (34, 37). Findings from earlier studies (7, 36, 38) informed and shaped the priority-setting process.

#### Workshop three

Workshop three focused on generating joint priorities and developing intervention options. Participants ranked priorities which the research team mapped to COM-B and the Theoretical Domains Framework (23, 26) (Table 1). Intervention ideas were generated and appraised using the APEASE framework (Acceptability, Practicality, Effectiveness, Affordability, Side effects, and Equity (23)). Following Workshop three, the research team coded the ideas to BCW intervention functions and BCTTv1 techniques (39) and developed a prototype rehabilitation programme.

**Table 1.** Joint priorities to address within the rehabilitation programme, mapped to the domains of COM-B and TDF.

| Joint priority to address within rehabilitation programme | COM-B domain | TDF domain |
| --- | --- | --- |
| 1. Improve patients' knowledge and understanding, and expectations of:<br>a. their condition<br>b. care processes<br>c. aims of surgery | Psychological capability | Knowledge |
|  | Reflective motivation | Beliefs about consequences |
| 2. Increase patients' perceived personal control of their condition and symptoms. | Reflective motivation | Beliefs about capabilities |
|  |  | Beliefs about consequences |
|  |  | Behavioural regulation |
| 3. Provide opportunity and access to pre-operative exercise to improve:<br>a. pre-operative walking capability and performance<br>b. lower limb strength<br>c. balance | Physical capability | Physical skills |
|  | Psychological capability | Knowledge |
|  | Physical opportunity | Environmental context and resources |
|  | Social opportunity | Social influences |
|  | Reflective motivation | Beliefs about capabilities<br>Beliefs about Consequences<br>Intentions |
| 4. Provide a personalised post-operative physiotherapy plan. | Physical opportunity | Environmental context and resources |
|  | Social opportunity | Social influences |
| 5. Address patient's fears, including fear of falling and moving. | Automatic motivation | Emotion |
|  | Physical capability | Physical skills |
| 6. Ensure timely communication for patients with surgical and rehabilitation team. | Physical opportunity | Environmental context and resources |
|  | Social opportunity | Social influences |
| COM-B: Capability, Opportunity, Motivation – Behaviour; TDF: theoretical Domains Framework |  |  |

#### Workshop four and small group meetings

In Workshop four the prototype programme was presented and refined. A modified Delphi process (40, 41) was used to address design uncertainties, with participants voting whether amendments were required for three components: inpatient physiotherapy, peer support, and supervised exercise. None reached the predefined 70% consensus threshold (40, 41), prompting further discussion on how the programme should be amended. The research team subsequently revised the rehabilitation programme. The updated prototype was reviewed in small group meetings (<30-minute, telephone or video call), with participants re-voting and achieving consensus. Content, format, and name were discussed and the final programme, named **STRIDE** (STructured Rehabilitation and InDividualised Exercise and education), was finalised following these consultations.

#### Activities conducted following the workshops and meetings

STRIDE programme materials and a clinician training manual were developed, informed by previous rehabilitation programmes (42). The patient advisory group reviewed the patient-facing materials, and made minor amendments to wording.

### The STRIDE programme

STRIDE is a 24-week physiotherapy led, behaviour change, rehabilitation programme delivered over 12 weeks before surgery (pre-operative phase) and 12 weeks after surgery (post-operative phase) (Table 2). Supplementary material 2 contains the logic model and TIDieR Checklist (31).

**Table 2.** Overview of STRIDE programme.

| Session | Format | Overview of session |
| --- | --- | --- |
| **Pre-operative phase. Duration 12 weeks** | 1<br>60 min<br>In person | • Address knowledge, understanding & expectations of surgery, care & outcomes<br>• Identification of personal concerns regarding condition, surgery, & outcome<br>• Determine baseline physical capability<br>• Instruct, demonstrate & practice balance, lower limb performance & flexibility exercises<br>• Provide & instruct on use of pedometer & walking diary<br>• Provide manual, signpost to educational videos |
|  | 2<br>30 min<br>Virtual or in person | • Review & provide feedback on walking & exercise diaries<br>• Establish walking goals & engage the participant in action planning to achieve them<br>• Discuss importance of & establish availability/plan for social support<br>• Modify exercise programme as required |
|  | 3<br>30 min<br>Virtual or in person | • Review & provide feedback on progress towards walking-based goals<br>• Prepare for surgery & peri-operative period including relevant advice on pain management, do & don'ts, any restrictions, & exercises in the immediate post-operative period<br>• Review & provide feedback on walking & exercise diaries, modify programmes as required |
|  | 4*<br>30 min<br>Virtual or in person | • Optional session<br>• Address specific concerns or problems e.g. progression of walking, balance, pain management, fear of movement etc.<br>• Review & provide feedback on progress towards walking-based goals<br>• Support the participant to progress & self-manage<br>• Discuss participant progress & problem solve issues |
| **Post-operative phase. Duration 12 weeks** | 1<br>30 min<br>Telephone | • Identify & address health & treatment concerns<br>• Provide reassurance<br>• Encourage resumption of walking & exercise programme |
|  | 2<br>30 min<br>In person | • Physical assessment including back & lower limb flexibility, lower limb performance & balance<br>• Review & provide feedback on progress towards walking-based goals<br>• Review & provide feedback on walking & exercise diaries, modify programmes as required |
|  | 3<br>30 min<br>Virtual or in person | • Review & provide feedback in relation to walking & exercise programme & progress relative to goals.<br>• Troubleshoot new problems<br>• Support planning for further self-management & maintenance of walking exercise behaviour<br>• Discharge the participant from STRIDE |
|  | 4 & 5*<br>30 min<br>Virtual or in person | • Optional sessions<br>• Address specific concerns or problems e.g. progression of walking, balance, pain management, fear of movement etc.<br>• Discuss participant progress & problem solve issues<br>• Support the participant to progress & self-manage |
| *Optional sessions; Virtual: telephone or video call dependent on patient preference |  |  |

#### Aim

The primary aim is to improve walking performance after surgery for LSS claudication

#### Format

The pre-operative phase includes three core sessions and one optional session; the post-operative phase includes three core sessions and up to two optional sessions. Two sessions are delivered in person in the physiotherapy clinic, the others may be conducted in person, or via telephone or video calls depending upon patient preference (Table 2).

#### Content

Sessions are tailored to address individual knowledge, expectations, perceived control, fears and concerns regarding their condition and treatment. Walking and exercise goals are collaboratively set. Sessions incorporate behaviour change techniques (BCTs) and promote self-management, engagement, and adherence to an individualised multi-modal exercise programme aimed at increasing walking behaviour, balance, lower limb strength and flexibility, and function. The STRIDE programme was coded to 15 BCTS, six intervention functions: *education, training, enablement, persuasion, modelling,* and *environmental restructuring*; and the policy category *service provision* from the BCW.

#### Resources

All patients receive a pedometer, personalised workbook (including information about LSS, their surgery, exercises and a walking and exercise diary). Participants also had access to online exercise and patient experience videos to support their preparation and recovery.

### Feasibility study

A multiple-methods, single-arm study of the co-developed rehabilitation programme, STRIDE was conducted. It comprised (i) quantitative assessment of feasibility outcomes and clinical outcomes, (ii) mixed-methods assessment of acceptability (questionnaires plus qualitative focus groups).

The aims were to assess the feasibility of conducting a future trial and acceptability of the STRIDE rehabilitation programme and to identify modifications to optimise delivery.

#### Setting and recruitment

Potentially eligible patients were identified from the surgical waiting lists at King’s College Hospital by one researcher.

### Eligibility criteria

#### Inclusion

aged ≥50 years; radiographic degenerative (LSS) with symptoms of neurogenic claudication; on a waiting list for elective lumbar decompression (with or without fixation); conversational English or willingness to use an interpreter.

#### Exclusion

LSS due to tumour or fracture; another primary condition limiting walking; inability/unwillingness to provide informed consent; lumbar fusion at >2 levels.

### Measurement and Procedures

Demographic, social and past medical history data were self-reported via questionnaires at baseline (T0) and included: age, body mass index (kg/m2), sex, ethnicity, education, employment, indices of deprivation, co-morbidities, and falls history.

### Feasibility, recruitment, and retention

The numbers of people approached and consented, retention at the end of the prehabilitation (T1) and post-operative (T2) phases, and questionnaire completion, adverse events, and attendance at STRIDE sessions were recorded. No a priori progression criteria were set; feasibility data guided modifications instead of progression decisions, though achieving the target sample size and median acceptability scores of ≥4 (out of 5) were considered indicators of feasibility.

### Clinical outcome measures

Outcomes were collected at baseline (T0), after the pre-operative phase (T1), and after the post-operative phase (T2) and included *walking capacity* (Six-Minute Walk Test, distance in metres (43)); *walking performance* (mean daily steps via triaxial thigh-worn accelerometer; ActivPal3™, PAL Technologies, Glasgow, UK); *perceived difficulty of walking* (0–10 numerical rating scale0 = no difficulty, 10 = unable to walk); *lower-limb function* (Five-Times Sit-to-Stand test (44) and Four-Stage Balance Test (45); *back and leg pain* (at rest and during walking, 0–10 (46)); *back-related disability* (Oswestry Disability Index, 0–100% (47)); *distress* (PHQ-4 (48)); *fear of falling* (Short Falls Efficacy Scale-International (49)); *fear of movement* (Tampa Scale of Kinesiophobia (50); *illness perceptions* (Brief IPQ (51)); *expectations* (modified version of the “expectations scale” of the North American Spine Society Lumbar Spine Questionnaire (52)); and c*hange in knowledge (*about condition, recovery, surgical/care processes, rated 0-10 scale studies (53). See Supplementary Materials 3.

### Fidelity of delivery

To assess delivery fidelity, the physiotherapist completed a session-specific checklist derived from the STRIDE manual, rating each item (fully/partially/not delivered) and recording session duration. We calculated global and item-level fidelity scores.

### Acceptability assessment

#### Questionnaires

At T0, T1, and T2 participants completed an acceptability questionnaire based on the Theoretical Framework of Acceptability (TFA) (54). The measure included 8-items on affective attitude, burden, opportunity costs, self-efficacy, intervention coherence, two perceived-effectiveness items, and a single overall acceptability item, all rated on 5-point Likert scales (1–5), with higher scores indicating greater acceptability. We calculated composite scores and analysed each item separately.

#### Focus groups

Two 90-minute audio-recorded focus groups were conducted (target ≤8 participants each). Group 1 (Microsoft Teams) focused primarily on capturing the acceptability of the pre-operative phase and was moderated by two researchers (CMc, HC) neither researchers were known to participants. Group 2 (in-person) focussed on the post-operative phase and was led by SMc with CMc support. Topic guides were developed a priori from the adapted TFA (55) and piloted with the patient advisory group (Supplementary materials 4). Data were transcribed verbatim, anonymised, and analysed thematically.

#### Procedures and schedule

After written informed consent, baseline assessments were completed (T0). The first STRIDE appointment was scheduled >1 week after baseline assessment to allow accelerometry wear. Objective assessments were conducted in person; questionnaires were completed on paper or electronically via Qualtrics (Qualtrics, Provo, UT), according to preference. Figure 2 illustrates the participant flow and study schedule.

**Figure 2.**
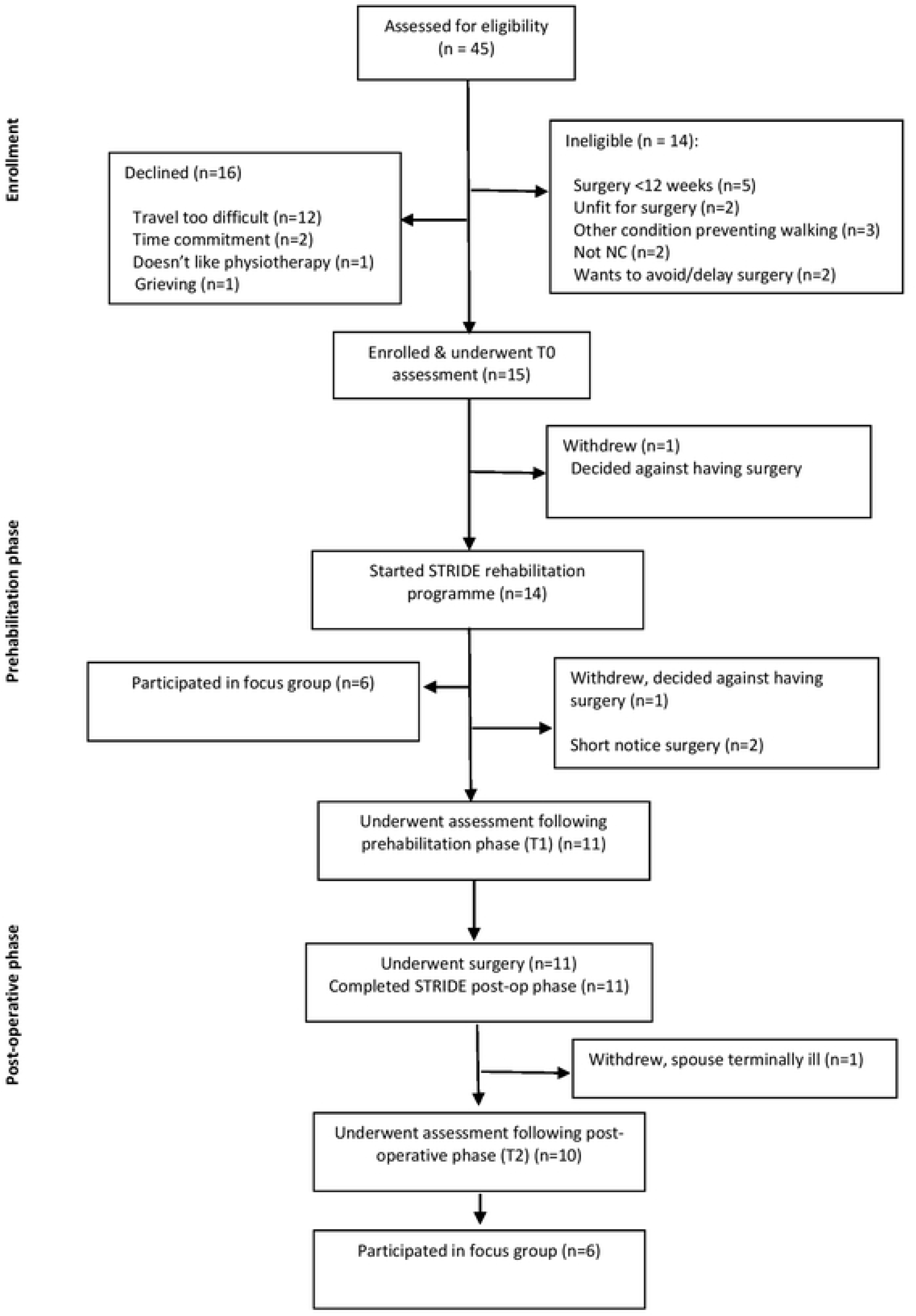
CONSORT flow of participants.

#### Programme delivery

All STRIDE sessions were delivered by SMc, a physiotherapist with >20 years’ experience and an MSc in Advanced Physiotherapy, who had completed short-course training in motivational interviewing. Delivery was manualised; proformas supported within-session content and documentation. After each session, SMc completed the fidelity checklist.

#### Sample size

As an acceptability study, no formal power calculation was performed. A target of n=15 was set based on guidance recommending minimum samples of 12 per arm for pilot trials (56) and balanced against timeframe constraints.

### Analysis

#### Quantitative analysis

Descriptive statistics summarised feasibility, acceptability, and clinical outcomes. Continuous data are reported as mean ± SD or median (IQR) as appropriate; categorical data as n (%). Clinical outcomes were summarised descriptively; standardised mean change effect sizes (Hedges’ g with small-sample correction) were calculated with 95% CIs using complete cases only (57). Analyses were conducted in Stata 18 (StataCorp, College Station, TX).

#### Qualitative analysis

Focus-group data were analysed using thematic analysis, combining deductive coding (mapping data to TFA constructs) with inductive coding to capture nuanced or unanticipated content (58). The process involved familiarisation, concurrent deductive and inductive coding, theme development within and across constructs, iterative refinement and naming, and synthesis for reporting. This approach offered a structured, theory-informed analysis while maintaining flexibility to incorporate emergent insights.

### Patient and public involvement and engagement

Service users, including patients, carers, and healthcare professionals, were central to this study. Patient advisors contributed to planning both studies, helping ensure the research was relevant, high quality, transparent, and acceptable. A patient advisory group (n=4, mixed gender and ethnicity) met with the research team at least every nine months and provided key input, including requesting that all patient workshops be held in person. Although unable to co-facilitate workshops due to availability or ill health, they reviewed and edited the STRIDE workbook and piloted and refined the acceptability questionnaire and focus group topic guide.

## Results

### Recruitment and Retention

Between 19^th^ December 2023 and 18^th^ January 2024, 45 patients were screened; 14 (39%) were ineligible and 16 (52%) declined, mainly due to travel requirements. Fifteen participants (33% of those screened; 48% of those eligible) enrolled. Four participants (27%) did not complete the full STRIDE programme. One participant withdrew before attending any STRIDE sessions and another withdrew after starting the programme but before the T1 assessment, both after deciding against surgery. Two other participants had not undergone surgery by study close and therefore did not receive the post-operative phase of STRIDE. One participant completed the full STRIDE programme but withdrew before the T2 assessment due to a spouse’s illness (Figure 2 – CONSORT flow diagram). Table 3 presents the baseline sociodemographic and clinical characteristics of the study participants. All participants underwent laminectomy (8 single-level, 2 two-level, 1 three-level). Median post-operative stay was 2 nights (IQR 4; range 1–20). One participant was later listed for lumbar fusion due to worsening spondylolisthesis.

**Table 3.**
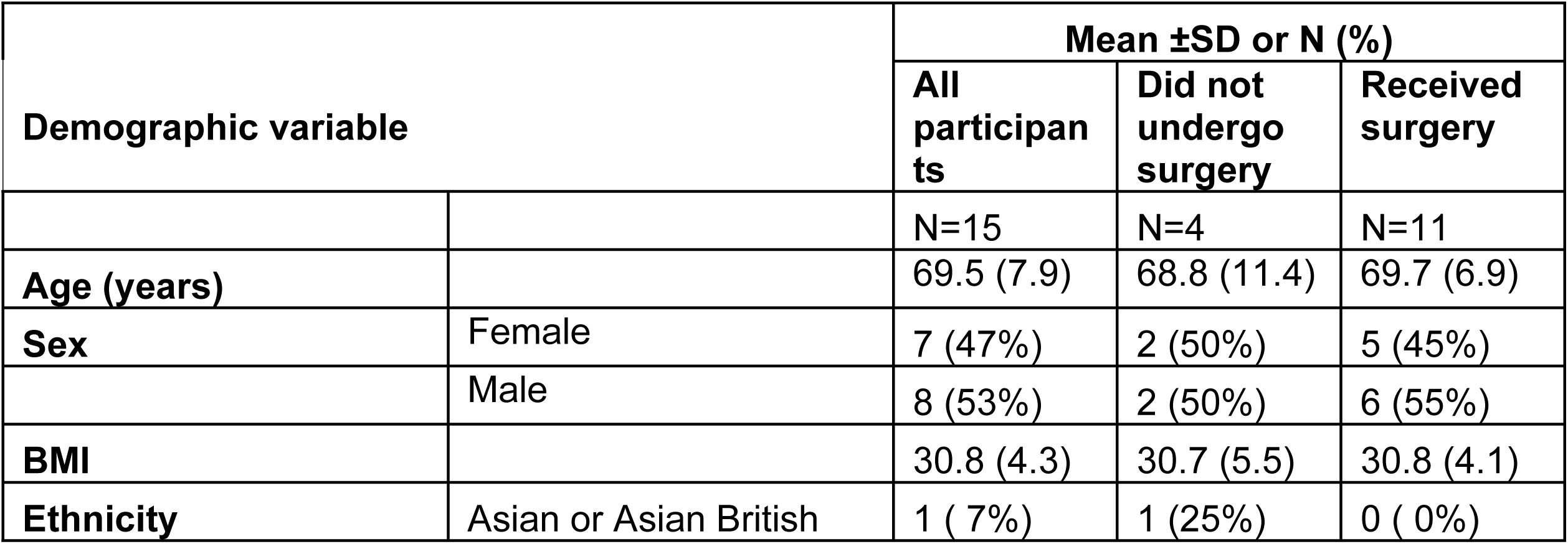
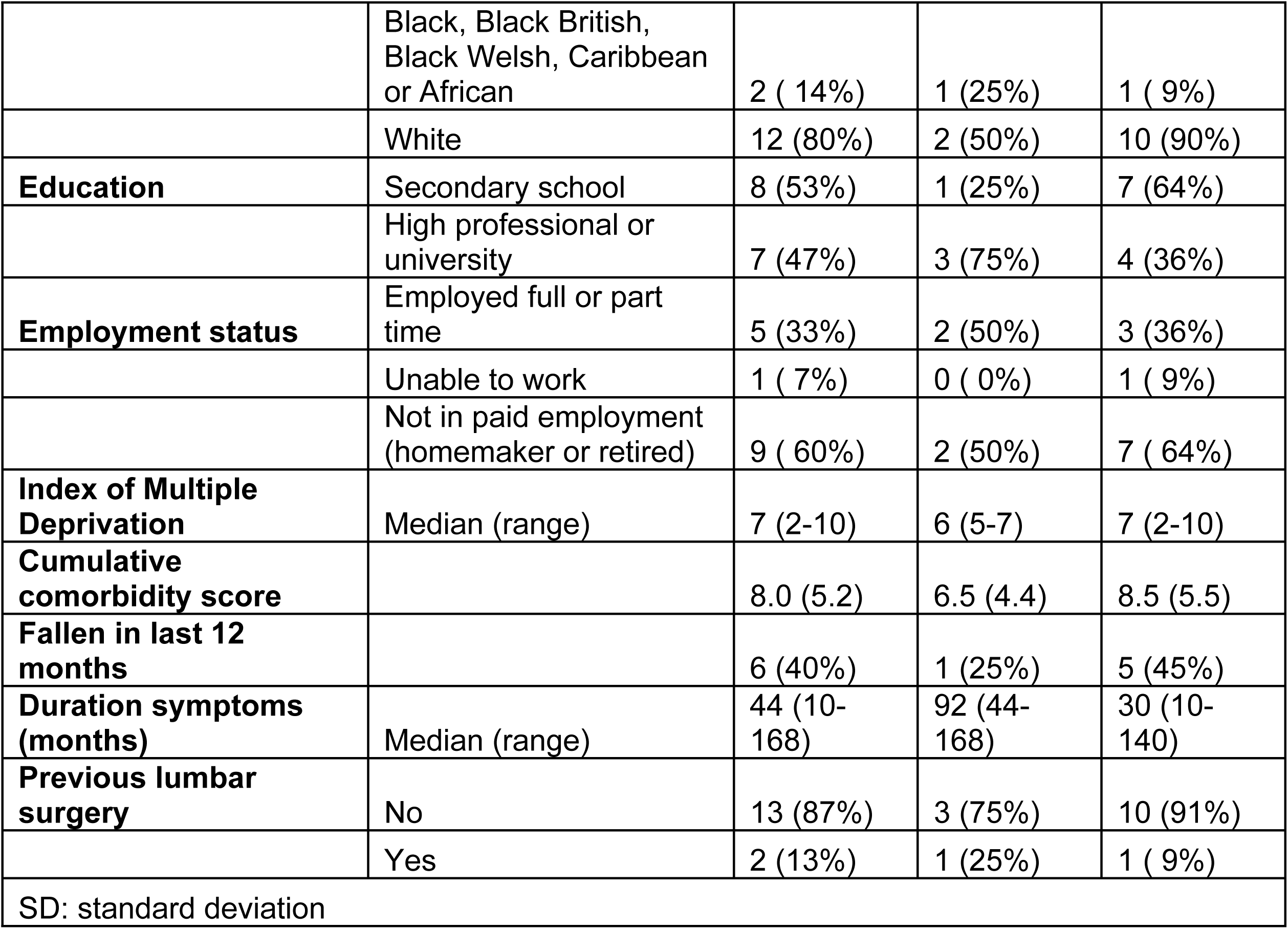
Feasibility study participant demographics.

#### Attendance and Adherence

STRIDE attendance was 82% for pre-operative sessions and 94% for post-operative sessions. Missed sessions were mostly due to short-notice surgery scheduling. Uptake of optional sessions was low (20% pre-operative; <10% post-operative). No adverse events were attributed to STRIDE.

#### Fidelity in the delivery of STRIDE

Delivery of core components was high: 91% were delivered, 7% part-delivered, and 3% omitted, usually due to time constraints. Additional non-manualised exercises were given to 64% (n=9) of participants. Two-thirds of sessions were delivered in set time targets; the second pre-operative and post-operative sessions often overran.

#### Clinical Outcomes

All 15 participants completed baseline assessments. At T1, data were available for only 11 (73%); however, only 4 (27%) completed all measures. This was largely due to short-notice surgery dates resulting in insufficient time for participants to attend for the objective assessment. At T2, 10 of 11 (91%) participants who had received surgery provided data. All participants provided valid accelerometer data meeting the minimum wear-time criterion (≥5 days), with a mean of 6.8 valid days per participant (median = 7; range = 5–7).

Table 4 presents the scores, change in scores and effect sizes for the walking, lower limb performance and disability outcomes; remaining outcomes are in Supplementary Materials 5. Outcomes generally showed small effect sizes at T1 and moderate to large effects at T2 assessments; but wide confidence intervals indicate low precision.

**Table 4.** Mean change in measures and effect sizes scores for the walking and disability outcomes.

| Variable | T0 mean (SD)<br>n=15 | T1 mean (SD)<br>n=7 | T2 mean (SD)<br>n=10 | T0-T1 mean<br>difference<br>(95%CI) | Effect size g<br>T0-T1 (95%CI) | T0-T2 mean<br>difference<br>(95%CI) | Effect size g<br>T0-T2 (95%CI) |
| --- | --- | --- | --- | --- | --- | --- | --- |
| 6MWD (m) | 288.6 (130.3) | 351.4 (157.6) | 361.0 (142.2) | 49.9 (27.1, 72.6) | 0.37 (-0.33, 1.08) | 81.6 (40.3, 123.0) | 0.63 (-0.08, 1.35) |
| Step count | 5459.5 (2798.6) | 6419 (2555.2) | 6526.8 (3809.7) | 868.4 (-424.7, 2161.5) | 0.3 (-0.40, 1.00) | 1405.1 (-53.5, 2863.6) | 0.5 (-0.21, 1.20) |
| 5 sit to stand (sec) | 19.2 (6.7) | 13.6 (5.8) | 13.8 (7.4) | -5.3 (-8.3, -2.2) | -0.83 (-1.55, -0.10) | -5.6 (-11.0, -0.2) | -0.88 (-1.61, -0.15) |
| 4-stage balance test | 3.7 (0.9) | 3.9 (1.3) | 4.2 (1.0) | 0.2 (-0.3, 0.6) | 0.21 (-0.48, 0.91) | 0.6 (-0.2, 1.3) | 0.68 (-0.04, 1.39) |
| Oswestry Disability Index | 40.7 (17.3) | 30 (20)* | 28 (24) | -3.1 (-9.0, 2.9) | -0.17 (-0.87, 0.52) | -14.1 (-23.6, -4.7) | -0.86 (-1.59, -0.13) |
| 6MWD: six-minute walk distance, CI: confidence intervals; *: n=8 |  |  |  |  |  |  |  |

#### Acceptability of STRIDE

Median overall acceptability was 5/5 (IQR 0), with 97% of responses in the highest categories. Constructs scoring the lowest were perceived burden and opportunity costs, reflecting challenges of travel and time demands (Figure 3).

**Figure 3.**
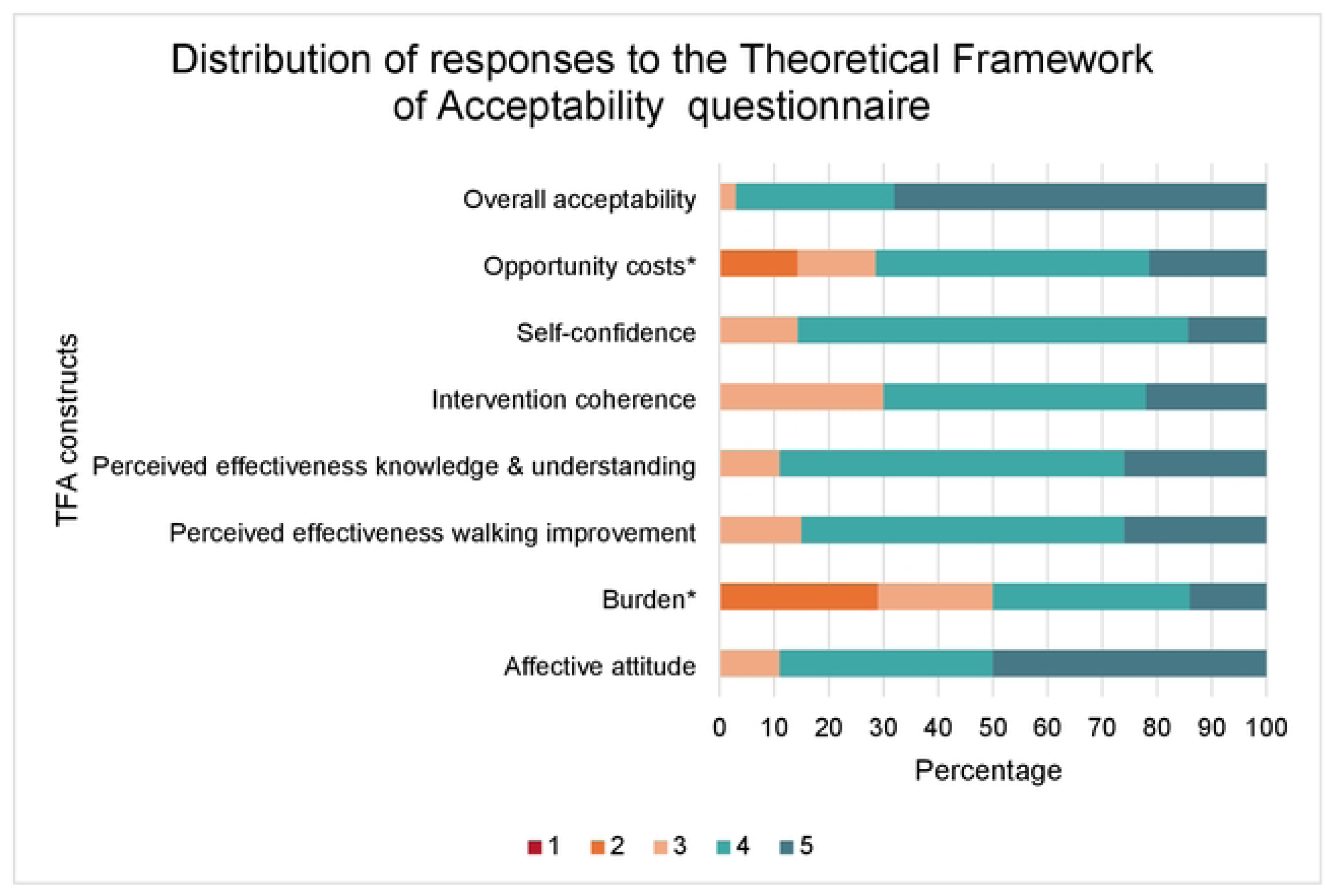
Distribution of responses to the TFA questionnaire. Responses rated on a Likert type scale 1-5, higher scores indicate greater acceptability. Burden and opportunity costs constructs have been reverse scored to ensure consistency in interpretation of scores.

### Focus groups

Seven participants attended the first pre-operative focus group, and eight participants attended the second post-operative focus group. In total, 12 unique participants took part, with three individuals attending both focus groups. STRIDE was described as highly acceptable. Participants emphasised the personalised approach, supportive physiotherapy input, and perceived effectiveness in increasing walking, and preparing for and recovering from surgery. Burden related to travel and time commitment represented the main challenges but were often deemed worthwhile given the programme’s benefits. Suggested refinements included improved resources, flexible delivery options, additional nutritional content, and structured peer support. Themes were generated aligning with all the constructs of the extended TFA. Table 5 provides a definition of each theme and participant-suggested refinements to enhance acceptability. Illustrative quotes are in Supplementary materials 6.

**Table 5.** Definition of themes aligned to extended constructs of the Theoretical Framework of Acceptability and recommendations to enhance acceptability.

| Theme | Definition | Recommendations to enhance acceptability |
| --- | --- | --- |
| TFA construct: Affective attitude |  |  |
| *STRIDE is a positive experience* | STRIDE was unanimously perceived as a beneficial and supportive experience that helped participants feel more active, confident, and reassured. Their positive experiences were shaped by enthusiasm, a sense of proactivity, emotional uplift, and an increased sense of well-being. | No recommendations identified |
| *The role of the supportive physiotherapist* | This theme underscores the value of the physiotherapist's supportive role in the STRIDE programme. By being a calm, knowledgeable, and reassuring presence, the physiotherapist was able to build trust, inspire confidence, and provide much-needed comfort to participants, making the programme more acceptable and effective from an emotional standpoint. | No recommendations identified |
| *Perceptions of supporting resources* | The supporting materials were seen as useful resources. The pedometer provided was inaccurate and many participants opted to source their own devices; some found the walking diaries cumbersome and the folder for the manual too bulky suggesting an online platform could improve ease of use. | • Revamp the walking & exercise diaries<br>• Manual available online<br>• Develop videos to make more engaging<br>• Offer reliable pedometer |
| TFA construct: Burden |  |  |
| *Navigating the Challenges of Attendance* | The primary challenge for participants was the effort required to travel to in-person appointments, especially due to walking difficulties. This suggests that reducing travel and offering flexible appointment times while maintaining care quality would alleviate this burden. | • Local provision<br>• Flexibility in appointment times and format<br>• Remote collection of objective measures |
| TFA construct: Ethicality |  |  |
| *The value of STRIDE* | Participants highly valued STRIDE's personalised approach, as it aligned with their values and needs by tailoring exercises to their individual capabilities, offering choice in information delivery and appointment formats, and giving them the autonomy to decide their level of participation. | • Ensure future participants have agency to decide whether they would like to participate in STRIDE and surgery is not contingent on this. |
| TFA construct: Intervention coherence |  |  |
| *STRIDE makes sense* | The sessions before surgery aimed at influencing post-operative outcomes made sense to participants. It was clear to participants that the content was aimed at preventing complications and promoting quicker recovery, with each appointment building on previous ones to maximise outcomes. | • Expanding content to include diet, nutrition, and mental health/ wellbeing. |
| TFA construct: Opportunity costs & gains |  |  |
| Worth the effort | Participants were willing to make sacrifices in terms of time and effort because they valued the benefits of the programme, both in terms of preparation and recovery from surgery, and continued support from healthcare providers. | No recommendations identified |
| TFA construct: Perceived effectiveness |  |  |
| *Promoting effective and sustained recovery* | STRIDE was perceived as effective in aiding preparation for and recovery from surgery, improving both physical and mental well-being, and providing long-term health benefits. | • Videos – greater range of experiences. |
| TFA construct: Safety and risks |  |  |
| *The safety of STRIDE* | Participants perceived the programme as safe, with minimal concerns expressed. | No recommendations identified |
| TFA construct: Self-efficacy |  |  |
| Fostering confidence to self-manage recovery | STRIDE was perceived to effectively enhance self-efficacy and motivation through personalised support, goal setting, and monitoring tools. Peer support could enhance motivation. | • Peer support groups<br>• Follow-up phone call<br>• Referral to local exercise group on completion of the programme |

## Discussion

An iterative, structured and theoretically informed approach was used to codesign STRIDE, a physiotherapy-led, behaviour change, rehabilitation programme. The feasibility study demonstrated that STRIDE was acceptable across multiple TFA domains to participants, with adequate recruitment and retention, and good attendance and engagement, supporting the feasibility of proceeding to a future trial. Some data-collection challenges were identified, alongside areas for modification of the programme and trial assessment procedures.

Participants reported that STRIDE supported surgical preparation and recovery. STRIDE was perceived as effective in improving walking performance, physical capability, and mental well-being. Suggested refinements included delivery at a local healthcare facility, incorporating peer support groups, and enhancing supporting materials. These findings inform refinements to intervention delivery and trial procedures ahead of a future trial.

STRIDE is novel because it provides a comprehensive programme targeting physical and psychosocial factors before and after surgery in people with LSS. Although some existing programmes span both phases, they typically emphasise pre-operative care (20, 59) or use cognitive-behavioural strategies without standardised post-operative rehabilitation (60, 61). STRIDE addresses these gaps with a balanced, integrated approach that may enhance short- to medium-term benefits observed in other programmes.

STRIDE aligns closely with recent evidence on spine prehabilitation. A realist review including 67 studies identified that effective prehabilitation requires clear communication, personalisation, accessibility and ongoing support (62), all of which were embedded within STRIDE’s design. An international consensus similarly prioritises education, psychological and physical preparation, multidisciplinary input and lifestyle factors (63). STRIDE operationalises these domains through structured education, behaviour-change strategies and progressive physical training.

STRIDE is distinguished by its explicit theoretical underpinning and comprehensive targeting of behavioural and physical determinants of walking. Grounded in the BCW, STRIDE conceptualises walking as a behaviour and systematically targets capability, opportunity and motivation, an approach shown to improve the effectiveness, transparency and replicability of behaviour change interventions (23). While a previous programme combined a walking programme with BCTs (16), STRIDE extends this approach by integrating progressive, supervised physical rehabilitation. Specifically, STRIDE targets lower-limb strength and balance alongside walking, key determinants of post-operative walking performance (7) and mobility in older adults (64, 65) that are underrepresented in rehabilitation programmes for people with LSS.

The integration of BCW and EBCD ensured meaningful patient involvement and a clear rationale for STRIDE’s mechanisms and content however, both required pragmatic adaptation. The EBCD discovery phase drew on multiple researcher-generated data sources, and the video component was omitted. While these deviations may limit methodological purity, the emotional mapping identified over 100 touchpoints, suggesting that core EBCD principles were retained. Tensions between experiential knowledge and theoretical rigour are recognised in co-design approaches (66, 67). Retrospective mapping of co-design outputs to BCW components was therefore a deliberate and pragmatic strategy. Although this diverges from the BCW’s prescribed sequence, it may enhance the practical relevance of theory-informed interventions while preserving fidelity to EBCD principles (68, 69).

In the feasibility study recruitment was 48% of those eligible, comparable to some lumbar surgery rehabilitation studies (16, 20, 60) but lower than others (70, 71). Travel requirements were the main barrier to participation and the greatest burden for those enrolled. Lower recruitment may reflect the older, mobility-restricted population and the pre-operative timing of STRIDE, when participants may feel less able to travel compared with post-operative studies where individuals are already experiencing the benefits of surgery e.g. pain reduction (72). Despite this, recruitment supports the feasibility of a future trial. Offering local clinical delivery sites therefore reducing travel demands may improve uptake, though barriers such as severe pain and low perceived value of prehabilitation may require targeted strategies.

Attrition was low among those who underwent surgery (91%) but higher in the whole study cohort (67%) due to participants either deciding against surgery or delays in surgery in the study timeframe. Outcome-measure completion at T1 was low due to surgery scheduled at short-notice and insufficient monitoring of electronic questionnaire completion. Remote monitoring (e.g., app-based walk tests) warrants exploration but may be unsuitable for those with severe walking limitation. Variable waiting-list times complicated the planning of prehabilitation. Potential solutions discussed by the focus group included a staged intervention (initial session upon joining the waiting list, followed by a more intensive phase closer to surgery), booster sessions, and peer support to sustain motivation, all require consideration.

STRIDE was highly acceptable to participants, perceived as safe and effective, as indicated by both quantitative and qualitative TFA data. Participants’ perceptions were supported by attendance and improvements in objective physical performance at T1 and T2. Burden was the lowest scored construct driven mainly by travel to in-person sessions. Interestingly, despite the availability of virtual options, most participants preferred in-person delivery, indicating a willingness to accept some burden. Effort related to behaviours within the programme, e.g. monitoring walking and exercises, was seldom mentioned as a concern, suggesting good acceptability.

Participants’ positive feelings toward STRIDE and their experiences preparing for and recovering from surgery contrast with previous studies (36). A key driver was the physiotherapist, whose support fostered trust, reassurance, and self-efficacy, factors known to enhance engagement and outcomes in LSS and LBP rehabilitation (73). Educational components likely provided cognitive reassurance, which is associated with reduced concerns and better outcomes (74). Personalisation was also highly valued, with tailored education, exercises, and goal setting enhancing motivation, confidence and acceptability.

## Strengths and limitations

This study has several strengths. Conceptualising walking as a behaviour provided a clear rationale for using behaviour change theory and enabled the co-design of an evidence-based rehabilitation programme. The TFA offered a comprehensive framework to assess acceptability through mixed-methods.

Several limitations should be acknowledged. The co-design workshops and one focus group were facilitated by the intervention developer, and this may have introduced bias. However, mitigation strategies were implemented to minimise the impact of this, including a consensus-building exercises and independent co-facilitators. The feasibility study was single-arm, single-site, and delivered by the intervention developer, limiting generalisability. Adherence to prescribed walking and exercise was unclear as participant diaries were not formally assessed.

## Conclusion

This paper has described the development and evaluation of STRIDE, a physiotherapy-led rehabilitation programme designed to improve walking after surgery for LSS. Using a theoretically informed, user-centred process that combined the BCW and EBCD, patients, family members, clinicians, and researchers collaboratively co-designed a programme targeting six shared priorities. The feasibility study demonstrated that STRIDE was highly acceptable to patients and positively shaped participants’ pre- and post-operative experiences. Recruitment and retention were encouraging, though travel to in-person appointments posed a barrier for some. Overall, STRIDE was highly acceptable to participants and key trail processes (e.g. recruitment and retention) were shown to be feasible, with encouraging signals of benefit, supporting progression to a future multi-site randomised controlled trial.

## Data Availability

Data will be available on the Kings Open Research Data System after acceptance

## Acknowledgements

The authors would like to thank all the participants in this study and the patient advisory group who gave their time and expertise with generosity included but not limited to: Aminul Ahmed, Riccardo Bario, Gary Chan, Christine Comer, Raquel Calado, Irene Dela Cruz, Emma Godfrey, Gordan Grahovac, Richard Griffin, Samuel Hatherley, Amy Jones, Chris Jukes, Nigel Kay, Margaret Kay, Trevor Morlese, Daniel Morris, Elayne Morris, Marion Mueller, Gary Pattinson, Anita Pattinson, Lawrence Quashie, Doreen Quashie, Geoff Sullivan, Joshua Tan, Victoria Udi, Laura Warne, Louise White, Julie Whitney, Dore Young, Constantine Zarifeh, Jackie Zarifeh.

## Supporting information captions

S1 Appendix. Codesign method and results

S2 Appendix. Logic model and TIDieR checklist for STRIDE

S3 Appendix. Clinical outcome measures within feasibility study

S4 Appendix. Focus group topic guides

S5 Appendix. Additional clinical outcomes

S6 Appendix. Illustrative quotes from focus groups

